# Targeting the centromedian thalamic nucleus for deep brain stimulation

**DOI:** 10.1101/19008136

**Authors:** Aaron E.L Warren, Linda J. Dalic, Wesley Thevathasan, Annie Roten, Kristian J. Bulluss, John S. Archer

## Abstract

**Objectives:** Deep brain stimulation (DBS) of the centromedian thalamic nucleus (CM) is an emerging treatment for multiple brain diseases, including the drug-resistant epilepsy Lennox-Gastaut syndrome (LGS). We aimed to improve neurosurgical targeting of the CM by (i) developing a structural MRI approach for CM visualisation, (ii) identifying the CM’s neurophysiological characteristics, and (iii) mapping connectivity from CM-DBS sites using functional MRI (fMRI).

**Methods:** Nineteen patients with LGS (mean age=28 years) underwent pre-surgical 3 tesla MRI using magnetisation-prepared <u>2</u> rapid acquisition gradient echoes (MP<u>2</u>RAGE) and fMRI sequences; 16 proceeded to bilateral CM-DBS implantation and intraoperative microelectrode recordings (MERs) from the thalamus. CM visualisation was achieved by highlighting intrathalamic borders on MP<u>2</u>RAGE using Sobel edge-detection. Mixed-effects analysis compared two MER features (spike firing rate, background noise) between ventrolateral, CM, and parafasicular nuclei. Resting-state fMRI connectivity was assessed using implanted CM-DBS electrode positions as regions-of-interest.

**Results:** The CM appeared as a hyperintense region bordering the comparatively hypointense pulvinar, mediodorsal, and parafasicular nuclei. At the group-level, reduced spike firing and background noise distinguished CM from the ventrolateral nucleus; however, these trends were not found in 20-25% of individual MER trajectories. Areas of fMRI connectivity included basal ganglia, brainstem, cerebellum, sensorimotor/premotor and limbic cortex.

**Conclusions:** In the largest clinical trial cohort of LGS patients undergoing CM-DBS reported to date, we show that accurate targeting of the CM is achievable using 3 tesla MP<u>2</u>RAGE MRI. MERs may provide additional localising features in some cases, however their utility is limited by inter-patient variability. Therapeutic effects of CM-DBS may be mediated via connectivity with brain networks that support diverse arousal, cognitive, and sensorimotor processes.

## INTRODUCTION

Deep brain stimulation(DBS) of the centromedian thalamic nucleus (CM; also termed céntre median nucleus) is an emerging treatment for diverse neurological and psychiatric diseases including epilepsy, Tourette syndrome, neuropathic pain, and disorders of consciousness[1].

The CM is a spheroid-shaped nucleus approximately 10mm in diameter that forms part of the posterior intralaminar thalamus together with the parafasicular (PF) and central lateral (CL) nuclei[1]. Evidence of its connectivity is limited in humans[2]; however, animal studies show connections with the basal ganglia (chiefly the striatum), brainstem, and select cortical regions including sensorimotor, premotor, and limbic cortex[3]. The CM’s connectivity is theorised to play a key role in a range of arousal, cognitive, and sensorimotor processes[1].

Lennox-Gastaut syndrome (LGS) is a severe form of epilepsy for which CM-DBS has shown potential efficacy. LGS typically begins in childhood and is characterised by multiple seizure types, generalised interictal epileptiform discharges on scalp electroencephalography (EEG), and cognitive and behavioural impairments that worsen over time[4]. Several placebo-controlled[5 6] and unblinded[2 7–9] trials of CM-DBS have been performed in LGS, with mixed results. Positive outcomes appear dependent on accurate positioning of electrodes within the CM[7].

Current opinion within the epilepsy DBS community is that the CM is not directly visualisable using structural neuroimaging[10]. Instead, pre-surgical targeting is typically nperformed indirectly[6 7 9], where a two-dimensional stereotactic atlas of the thalamus, such as the one published by Schaltenbrand and Wahren in the 1970s[11], is superimposed on a CT or MRI scan relative to coarse anatomical landmarks including anterior and posterior commissures.

However, indirect targeting poorly accommodates inter-patient variability in thalamic anatomy, which can lead to inaccurate placement of DBS electrodes, suboptimal therapeutic response, and increased frequency of surgical complications and adverse side-effects[12]. These risks are of particular concern in LGS, given that up to 50% of LGS patients have structural brain abnormalities[13], with their neuroanatomy likely deviating from stereotactic-based atlases.

In DBS surgery for other diseases, targeting is commonly aided by microelectrode recordings (MERs), where electrode positions are refined intraoperatively according to neurophysiological ‘signatures’ of the stimulation target. For example, in Parkinson’s disease, MERs show that the subthalamic nucleus is distinguishable from surrounding structures by changes in neuronal spike firing and background noise[14]. Comparatively few studies have assessed the utility of MERs in localising stimulation targets for epilepsy[15].

We aimed to (i) develop a structural MRI approach for pre-surgical visualisation of the CM in LGS; (ii) assess whether MERs assist with intraoperative identification of the CM; and (iii) investigate functional connectivity from CM-DBS sites using resting-state fMRI.

## MATERIALS AND METHODS

### A note on brevity

Detailed documentation of materials and methods is in Supplementary Material. An abridged version highlighting key methodological details only is provided here.

### Patients

Nineteen patients with LGS (mean age=28 years) were recruited for the Electrical Stimulation of the Thalamus for Epilepsy of Lennox-Gastaut Phenotype (ESTEL) study: a randomised, double-blind, placebo-controlled clinical trial of CM-DBS at Austin Health in Melbourne, Australia (Australian Department of Health, clinical trial notification CT-2016-CTN-05306-1; Human Research Ethics Committee approval number HREC/16/Austin/139). Written informed consent was obtained from each patient’s parent/guardian.

### Pre-surgical MRI

Pre-surgical MRI was acquired in 3 tesla Siemens Skyra scanners. Eight patients were scanned using a Siemens transmit-receive (Tx/Rx) CP head coil; seven using a Siemens 64-channel head/neck coil; and four using a Siemens 32-channel head/neck coil. In all, we acquired a magnetisation-prepared <u>2</u> rapid acquisition gradient echoes (MP<u>2</u>RAGE) sequence[16], and a magnetisation-prepared rapid gradient echo (MPRAGE) sequence with intravenous gadolinium contrast; the latter served as the primary reference for neurosurgical trajectory planning. MP<u>2</u>RAGE differs from MPRAGE by acquiring two volumes with different inversion times which are combined into a unified volume that shows improved grey/white matter image contrast[16]. In 14 patients only, resting-state fMRI was also acquired in a separate pre-surgical scan.

### MP2RAGE processing for CM visualisation

An edge-detection method (Sobel operator) was used to highlight intrathalamic borders visible on MP<u>2</u>RAGE. The Sobel operator computes the gradient magnitude at each image point (i.e., change in signal intensity from one voxel to the next), where areas of high gradient indicate likely ‘edges’. Voxel intensities within each of the MP<u>2</u>RAGE and Sobel-processed images were converted to *z-*scores, and then a novel ‘edge-weighted MP<u>2</u>RAGE’ image was calculated by summing together the two *z*-score images: this weighted each MP<u>2</u>RAGE voxel by its likelihood of indicating a border between areas of differing signal intensity (i.e., border voxels became ‘brighter’ in the summed image, whereas non-border voxels became ‘darker’).

### Comparison with three-dimensional thalamic atlases

We compared the CM’s location on edge-weighted MP<u>2</u>RAGE to two three-dimensional atlases in asymmetric Montreal Neurological Institute (MNI)152 2009b template space (Figure 1): the ‘Krauth/Morel’ atlas[17], which represents a digital mean across six specimens of the human thalamus and is available as a set of 72 binary masks; and the ‘BigBrain’ atlas[18], which displays histology of a complete human brain. Comparisons were achieved by calculating nonlinear spatial warps between patients’ MP<u>2</u>RAGE scans and MNI space.

**Figure 1:**
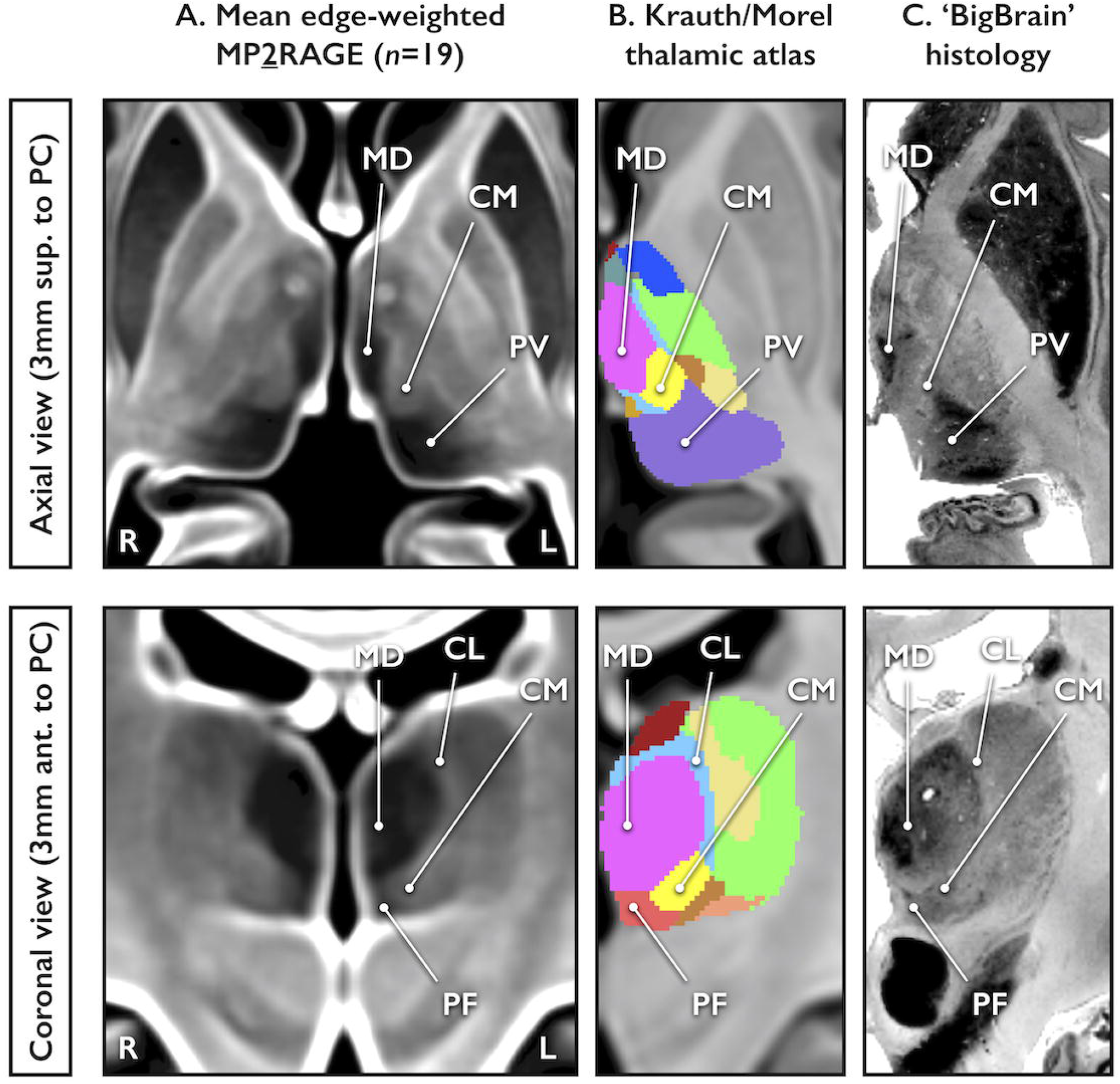
Group-level visualisation of centromedian nucleus on mean edge-weighted MP<u>2</u>RAGE and comparison with thalamic atlases. **(A)** Mean image computed across all patients’ (*n*=19) pre-surgical edge-weighted MP<u>2</u>RAGE scans following spatial warping to Montreal Neurological Institute (MNI)152 2009b template space. Axially (upper row), the centromedian nucleus (CM) appears as a hyperintense region bordered medially by the comparatively hypointense mediodorsal nucleus (MD) and posteriorly by the hypointense pulvinar nucleus (PV). Coronally (lower row), the CM is positioned inferior to the MD and medial to the hypointense parafasicular nucleus (PF), while the CM’s approximate lateral extent is suggested by a thin line of hyperintensity consistent with the centrolateral nucleus (CL). It is best appreciated ∼3 mm superior and ∼6 mm anterior to the posterior commissure (PC). **(B-C)** Atlas comparisons. ***Interpretation:*** Location of the CM on edge-weighted MP<u>2</u>RAGE showed good correspondence with the CM’s position determined by ‘Krauth/Morel’[17] and ‘BigBrain’[18] atlases.

**Figure 2:**
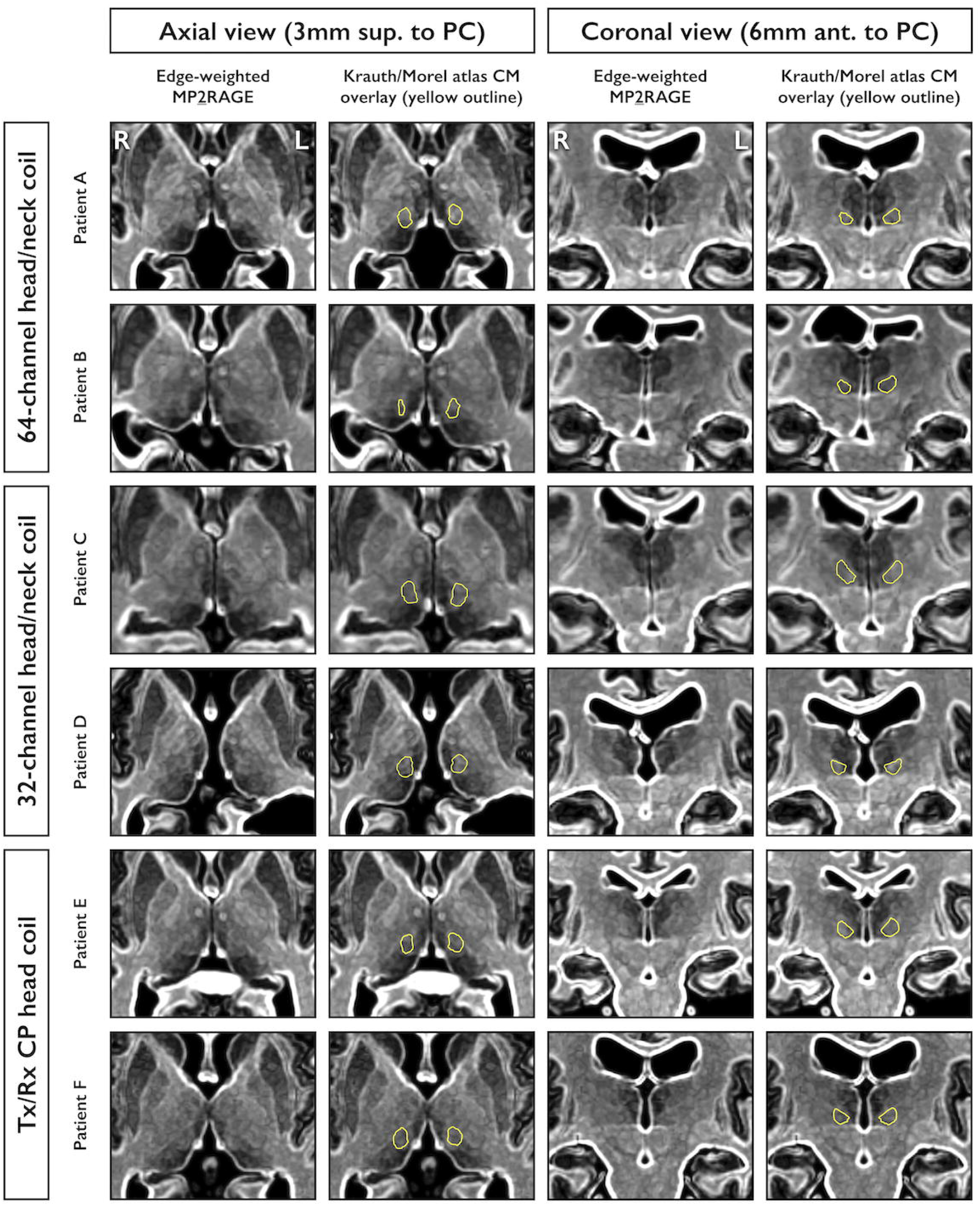
Individual-level visualisation of centromedian nucleus on edge-weighted MP<u>2</u>RAGE. Visualisation is shown for six example patients scanned using one of three different Siemens MRI head coils (two patient examples are shown for each of the 64 channel head/neck, 32-channel head/neck, and transmit/receive [Tx/Rx] CP head coils). Consistent with the group-level mean image (Figure 1), the centromedian nucleus (CM) appears as a hyperintense region and is best appreciated on axial views (far left two columns) ∼3 mm superior to the posterior commissure (PC), and on coronal views (far right two columns) ∼6 mm anterior to the PC. To assist identification, a binary mask of the CM from the Krauth/Morel atlas[17] has been nonlinearly spatially warped from Montreal Neurological Institute space to each patient’s own brain. Outlines of the resulting patient-space CM masks are displayed for each image in yellow.

### fMRI connectivity from CM-DBS sites

For each patient, whole-brain fMRI connectivity was calculated from two spherical regions-of-interest (ROIs; radius=3mm; one per hemisphere) centred on the DBS electrode contacts selected for monopolar stimulation (in most patients, the second-most inferior contact [i.e., contact 1] was selected). MNI coordinates of each ROI are provided in Table 1. For left- and right-hemisphere ROIs separately, we assessed whether mean connectivity strength across patients at each voxel was different from zero using non-parametric one-sample *t-*tests[19], with significance assessed at *p<*0.005 (family-wise error corrected) following threshold-free cluster enhancement[19]. *ICN_Atlas* software[20] was used to identify the ‘BrainMap’ intrinsic connectivity networks (ICNs) with which CM-DBS sites showed maximal functional connectivity[21].

**Table 1:** Coordinates of implanted DBS electrode contact positions for all patients. For each patient (*n=*16), three-dimensional coordinates (*x, y, z* [mm]) are provided in Montreal Neurological Institute (MNI)152 2009b template space for the four DBS electrode contacts on the left brain side (L C0, L C1, L C2, L C3), and the four contacts on the right (R C0, R C1, R C2, R C3). The mean coordinates (across all patients) are provided for each contact at the table’s bottom. Boxes shaded in yellow indicate electrode contacts located within the centromedian nucleus (CM), as determined by the Krauth/Morel atlas[17], while boxes additionally enclosed by a red border indicate electrode contacts that were selected for monopolar stimulation and were used in the resting-state fMRI connectivity analysis. *=these patients were not included in the fMRI analysis.

| Patient ID | Left DBS Contacts<br>MNI coordinates (x, y, z [mm]) |  |  |  | Right DBS Contacts<br>MNI Coordinates (x, y, z [mm]) |  |  |  |
| --- | --- | --- | --- | --- | --- | --- | --- | --- |
|  | L C0 | L C1 | L C2 | L C3 | R C0 | R C1 | R C2 | R C3 |
| 1 | -11, -20, 1 | -12, -18, 3 | -13, -18, 5 | -14, -16, 6 | 8, -17, 1 | 9, -15, 2 | 11, -14, 3 | 13, -14, 4 |
| 2 | -12, -24, -2 | -12, -23, -1 | -13, -21, 0 | -14, -19, 2 | 8, -25, -2 | 9, -23, -1 | 10, -22, 0 | 11, -20, 2 |
| 3 | -8, -20, -2 | -8, -19, 0 | -9, -18, 2 | -10, -16, 4 | 9, -20, -4 | 10, -19, -3 | 11, -17, -1 | 12, -16, 1 |
| 4 | -7, -21, -2 | -9, -20, -1 | -9, -19, 0 | -10, -18, 1 | 9, -21, -3 | 10, -19, -2 | 11, -18, 0 | 12, -17, 1 |
| 5 | -7, -19, -2 | -9, -18, 0 | -10, -18, 2 | -11, -17, 4 | 9, -21, -1 | 10, -21, 1 | 11, -21, 3 | 13, -20, 4 |
| 6 | -9, -22, -2 | -10, -21, -1 | -11, -19, 1 | -12, -18, 3 | 10, -22, -1 | 11, -20, 1 | 12, -18, 2 | 13, -17, 3 |
| 7 | -11, -21, 1 | -12, -20, 3 | -13, -18, 4 | -14, -17, 6 | 11, -21, 0 | 12, -19, 2 | 13, -18, 3 | 14, -16, 5 |
| 8 | -10, -22, 0 | -11, -20, 1 | -12, -18, 3 | -13, -16, 4 | 8, -21, -1 | 9, -19, 1 | 10, -17, 3 | 11, -15, 4 |
| 9 | -8, -20, -1 | -9, -19, 1 | -10, -18, 3 | -11, -17, 4 | 9, -21, 0 | 10, -18, 1 | 11, -16, 2 | 12, -14, 3 |
| 10 | -10, -22, 0 | -11, -21, 2 | -12, -19, 3 | -13, -18, 5 | 9, -21, -3 | 10, -20, -1 | 11, -18, 1 | 12, -17, 3 |
| 11 | -10, -22, -2 | -10, -21, 0 | -11, -19, 2 | -12, -17, 3 | 11, -22, -1 | 12, -20, 0 | 14, -19, 2 | 15, -17, 3 |
| 12 | -10, -21, -2 | -10, -20, -1 | -11, -19, 0 | -12, -17, 2 | 9, -20, 0 | 10, -19, 1 | 10, -18, 3 | 11, -17, 4 |
| 13 | -10, -19, -1 | -11, -17, 1 | -13, -15, 3 | -14, -14, 5 | 10, -19, 0 | 11, -18, 2 | 12, -16, 4 | 14, -15, 5 |
| 14 | -10, -21, 1 | -11, -20, 2 | -12, -18, 3 | -13, -16, 4 | 9, -22, -1 | 10, -21, 0 | 11, -19, 1 | 12, -17, 2 |
| 15* | -11, -24, 1 | -12, -22, 2 | -13, -20, 3 | -13, -18, 4 | 8, -22, 0 | 9, -21, 1 | 10, -20, 3 | 11, -18, 4 |
| 16* | -10, -23, -1 | -11, -22, 0 | -12, -20, 2 | -13, -19, 4 | 9, -21, -1 | 9, -20, 1 | 10, -18, 2 | 11, -17, 3 |
| Mean | -10, -21, -1 | -11, -20, 1 | -12, -19, 2 | -12, -17, 4 | 9, -21, -1 | 10, -20, 0 | 11, -18, 2 | 12, -17, 3 |

### DBS and post-surgical reconstruction of lead trajectories

Sixteen patients proceeded to bilateral implantation of quadripolar DBS leads (Medtronic 3389). Surgical planning was performed using a StealthStation navigation system (Medtronic, Ireland). Implantations (and thus also MERs) were performed under anaesthesia maintained using combined isoflurane (0.5-0.7% end-tidal concentration) and remifentanil (0.1-0.3mcg/kg/min).

The typical trajectory entered the thalamus at the level of the ventrolateral nucleus (VL), passed through the CL and CM, and terminated near the CM/PF border (Figure 3). CM target selection used a three-step approach: (i) initial estimation based on established coordinates[7 9 11]; (ii) estimation of CM outline using the Krauth/Morel atlas[17] nonlinearly warped to the patient’s brain; and (iii) patient-specific direct targeting of CM by identifying a hyperintense region on the edge-weighted MP<u>2</u>RAGE image consistent with locations determined by (i) and (ii). This allowed us to position electrodes such that 2-3 of the 4 contacts were typically inside CM (Table 1).

**Figure 3:**
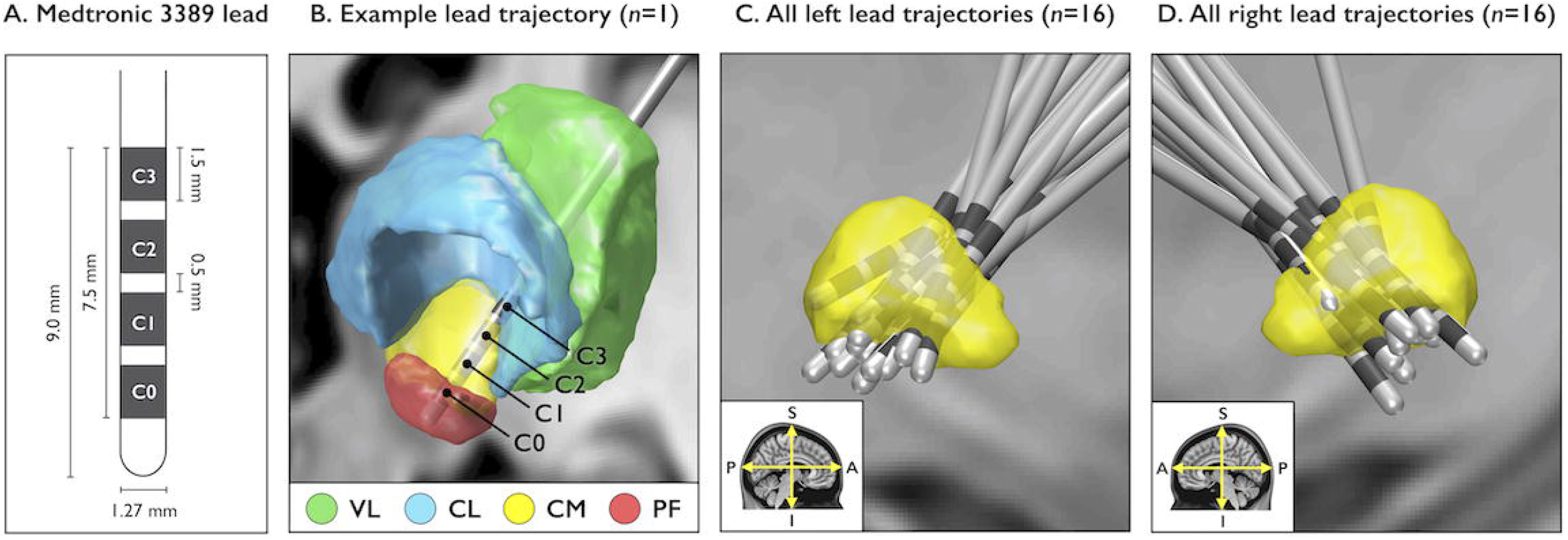
Positions of quadripolar DBS electrode leads (Medtronic 3389) bilaterally implanted into the centromedian nucleus. **(A)** Schematic showing lead width, contact size, and inter-contact distance for the Medtronic 3389 model. Each lead has four stimulation contacts (C0, C1, C2, and C3) spaced 0.5 mm apart. **(B)** Example reconstruction (from post-surgical CT; *n=*1) showing the typical lead trajectory and anatomical positions of contacts within thalamic nuclei (defined by the Krauth/Morel atlas)[17]. The typical trajectory positioned at least 2-3 contacts within the centromedian nucleus (CM), with the most inferior contact positioned near the border between the CM and parafasicular nucleus (PF). **(C-D)** Bilateral lead reconstructions for all patients (*n*=16), shown separately for the left and right CM (shapes in yellow). A sagittal view is provided for each brain side, as indicated by the directional arrows in the bottom left corner of each image (A=anterior direction; P=posterior direction; S=superior direction; I=inferior direction).

Post-surgical CT scans were used to confirm accuracy of DBS electrode positions. Lead trajectories were reconstructed from CT using lead-DBS software v2.1.8.1[22]. To visualise electrode positions in a common space (Figure 3), reconstructed leads were warped to MNI space. MNI coordinates of all patients’ DBS electrode contacts are provided in Table 1.

### Intraoperative MERs

Single-channel MERs were acquired with a LeadPoint system (Medtronic, Ireland). MERs were performed along 32 surgical trajectories from 16 patients, yielding 410 recording sites. Average recording duration across individual sites was 7s (range=3-12s). An average of 13 MERs were acquired per trajectory, typically beginning 10-12mm above the target and advancing in 1mm steps until reaching a depth of 1-3mm below target. MERs’ anatomical locations were determined by plotting their depths in mm (with respect to the target) along the CT-reconstructed lead trajectories[22], and then assigning them to the closest nucleus defined by the Krauth/Morel thalamic atlas[17].

Most MERs (361/410) were located in three nuclei sampled along the trajectory: 135 were in VL, 170 in CM, and 56 in PF. We excluded the remaining MERs from further analysis because they were located in other nuclei that each contained <5% of the total number of MERs. Three-dimensional coordinates of all included MERs are plotted in Figure 4A.

**Figure 4:**
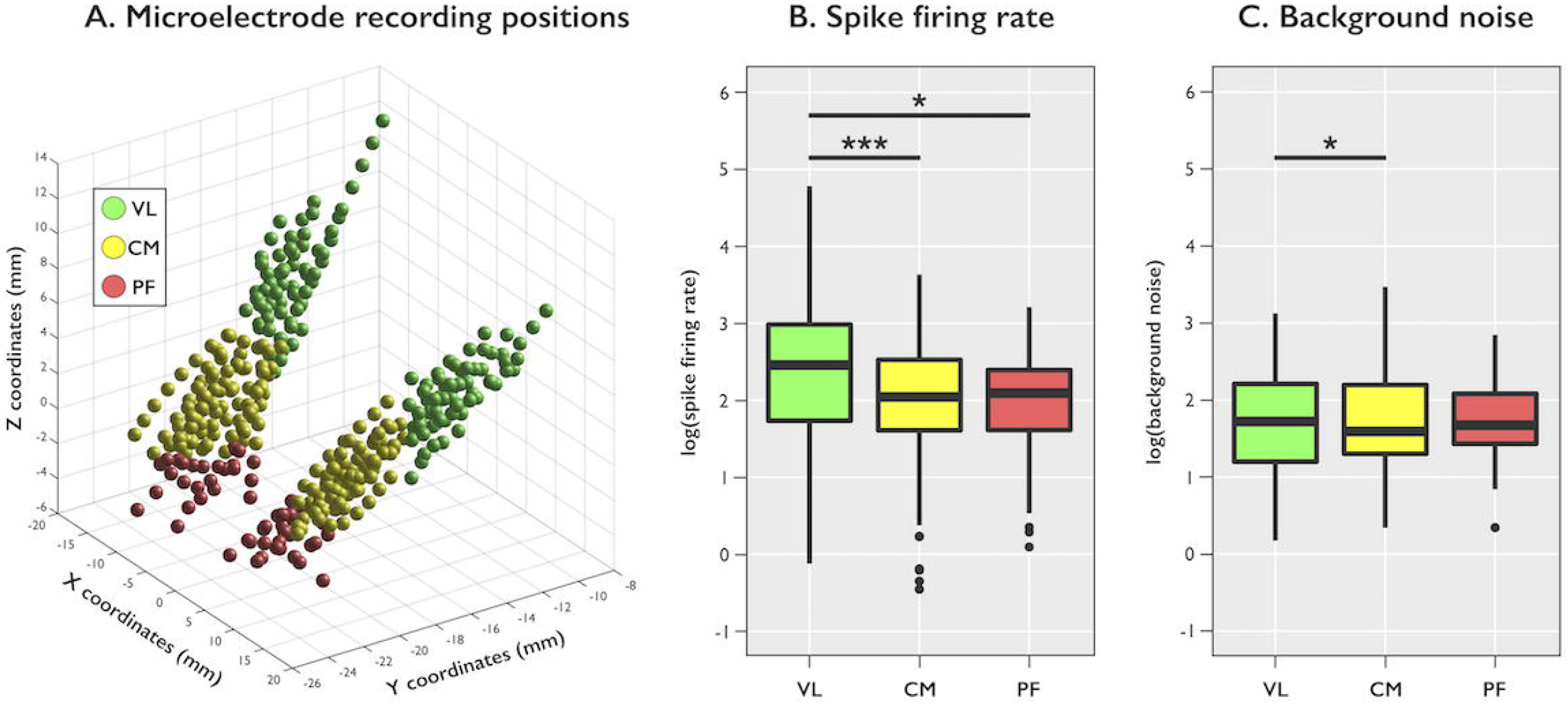
Group-level analysis of intraoperative microelectrode recordings. **(A)** Three-dimensional positions of the total 361 recording sites analysed from 16 patients. Recording sites are coloured according to their anatomical location defined by the Krauth/Morel atlas[17] (VL=ventrolateral nucleus; CM=centromedian nucleus; PF=parafasicular nucleus). Positions are displayed as *x, y*, and *z* coordinates (mm) in Montreal Neurological Institute (MNI)152 2009b template space. **(B-C)** Box plots showing post-hoc pairwise comparisons of spike firing rate and background noise (expressed as log values) between the VL, CM, and PF. The lower and upper ends of each box correspond to the first and third quartiles (i.e., spanning the inter-quartile range [IQR]) of recording values, respectively, while the middle horizontal line indicates the median. The lower and upper whiskers extend to the smallest and largest values no further than 1.5 times the IQR below the first quartile and 1.5 times the IQR above the third quartile. Outliers, defined as individual recording values beyond the whisker ends, are displayed as black circles. ***Interpretation:*** Both the CM and PF showed reduced spike firing rate compared to the VL, while background noise was lower in the CM compared to the VL (***=*p*<0.0001; *=*p<*0.01).

### Comparison of MER features between thalamic nuclei

MERs were analysed offline to assess whether they differentiate the VL, CM, and PF. For each MER, we computed *background noise* (using the mode of the signal envelope approach[23]) and *spike firing rate* (using the ratio between the number of spikes and the MER length in seconds[15]).

Using *R* v3.4.4, we performed group-level mixed-effects analyses to assess whether MER features varied by thalamic nucleus while accounting for variance due to MER sampling from different patients. Analyses were performed on log-transformed values of MER features. Type-II Wald *F*-tests tested for a main effect of nucleus, with significance assessed at *p*<0.05 (Bonferroni-corrected). Post-hoc tests were then used to compare each pair of nuclei via their estimated marginal means, with significance assessed at *p*<0.05 (Tukey-corrected).

To explore whether group-level results were seen at the individual level, we plotted spike firing rate and background noise for each MER trajectory separately. We only plotted trajectories where all three nuclei of interest were sampled and where representative spiking activity (≥5 spikes) was observed for at least one MER site per nucleus: thus 24 of the total 32 trajectories were plotted.

### Intraoperative microstimulation

After MERs, the microelectrode tip was withdrawn to target depth and stimulation was applied (130Hz, 60μs, 1-6mA) to determine the stimulation threshold at which clinical motor effects were elicited, reflecting the tip’s position with respect to the internal capsule[24]. No patients had any clinically detectable movement or change in muscle tone at <4mA current. At 4-6mA, subtle facial contractions or changes in contralateral limb tone were observed for three of the 32 target locations. Twelve target locations were additionally stimulated at higher currents (9-10mA), and similar motor effects were observed in seven. No target positions were adjusted as a result of these test stimulations.

## RESULTS

### CM visualisation on edge-weighted MP2RAGE

Orthogonal views of edge-weighted MP<u>2</u>RAGE are provided for six example patients in Figure 2, and the average across all 19 patients (after warping to MNI space) is displayed in Figure 1. The CM appeared as a hyperintense region, and on axial views its medial and posterior boundaries were demarcated by two comparatively hypointense regions consistent with the mediodorsal and pulvinar nuclei. Coronally, its medial and superior boundaries were identifiable by two hypointense areas consistent with the PF and mediodorsal nuclei. Its lateral boundary was less clearly defined, however on select coronal views a thin descending line of hyperintensity, consistent with the myelin-rich internal medullary laminae and CL, suggested an approximate lateral extent. The CM’s location on edge-weighted MP<u>2</u>RAGE visually corresponded with its position determined by Krauth/Morel[17] and BigBrain[18] atlases. Visualisation was possible across patients scanned using 64-channel, 32-channel, and Tx/Rx CP head coils.

Across all trajectories (left and right combined), the target coordinates were an average distance, with respect to the mid-commissural point determined in each patient’s native brain space, of 8.1mm lateral (range=6.7-10.6), 9.8mm posterior (range=2.4-12.6), and 1.1mm superior (range=-4-2.9); and the trajectory had an average rotational angle of 30.5° from the mid-sagittal plane (range=18.8-42.1) and 50.5° from the axial anterior commissure-posterior commissure(AC-PC) plane (range=35.3-70.7).

In several patients, our method for CM targeting clearly outperformed the ‘Schaltenbrand and Wahren’ stereotactic atlas-based method[7 9 11], where a two-dimensional atlas is superimposed (without nonlinear deformation) on the patient’s scan with respect to AC-PC landmarks. In particular, for patients with structural abnormalities (e.g., malformations of cortical development), the Schaltenbrand and Wahren atlas was grossly inaccurate for thalamic targeting (Figure 5).

**Figure 5:**
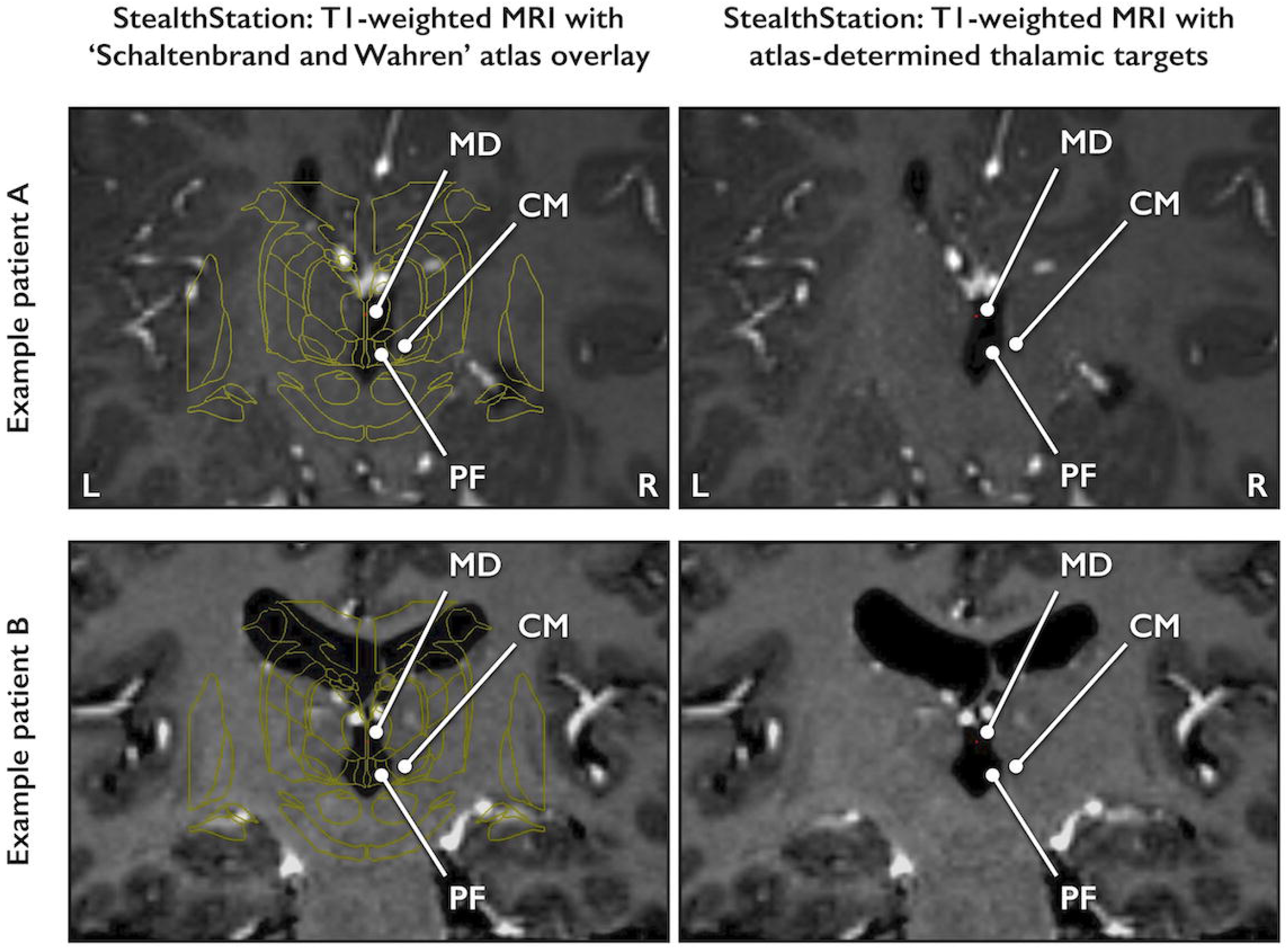
Patient examples illustrating potential inaccuracy of the indirect ‘Schaltenbrand and Wahren’ targeting method for identifying the centromedian nucleus. Example images are shown for two patients with LGS (one in each of row) who have structural brain abnormalities due to malformations of cortical development. The left column shows a coronal view of the two-dimensional ‘Schaltenbrand and Wahren’ thalamic atlas (plate 28)[11] superimposed on the patient’s T1-weighted MPRAGE MRI with respect to the anterior and posterior commissures, as performed using a StealthStation surgical navigation system (Medtronic, Ireland). White lines indicate the positions of the centromedian (CM), mediodorsal (MD), and parafasicular (PF) nuclei, as determined by the atlas. The right column shows the same coronal view of the patient’s MRI without the atlas overlaid but retaining the atlas-determined positions of CM, MD, and PF. ***Interpretation:*** these example patients’ thalamic anatomy shows significant deviation from the Schaltenbrand and Wahren stereotactic atlas. Indirect targeting based on this method alone would likely lead to misplacement of deep brain stimulation electrodes outside thalamic targets (e.g., in the examples shown, some atlas-determined nuclei locations are within cerebrospinal fluid).

### Intraoperative MER features

At the group-level, a main effect of nucleus was found for both spike firing rate(*F*_2,306_ =10.7, *p*<0.0001) and background noise(*F*_2,344_=4.9, *p<*0.01). Post-hoc tests (Figure 4B-C) showed that spike firing rate was lower within CM relative to VL(*t*_310_*=*4.2, *p*<0.0001), and within PF relative to VL(*t*_307_=3.5, *p*<0.01), but no difference was found between CM and PF(*t*_303_=0.4, *p*>0.9). Background noise was also lower within CM relative to VL(*t*_345_=3.1, *p*<0.01), but no difference was found between VL and PF(*t*_345_=1.8, *p*>0.1) nor between CM and PF(*t*_344_*=*0.5, *p*>0.8).

However, analysis of individual MER trajectories revealed some variability (Figure 6). Consistent with the whole-group results above, 80% of the individual trajectories showed lower mean spike firing within CM relative to VL; 80% showed lower mean spike firing within PF relative to VL; and 75% showed lower mean background noise within CM relative to VL. However, the remaining 20-25% did not follow these group trends. Relative differences between nuclei also showed variability: in some cases, there was a large difference in mean values between the VL and CM and between the VL and PF, whereas differences in others were more subtle.

**Figure 6:**
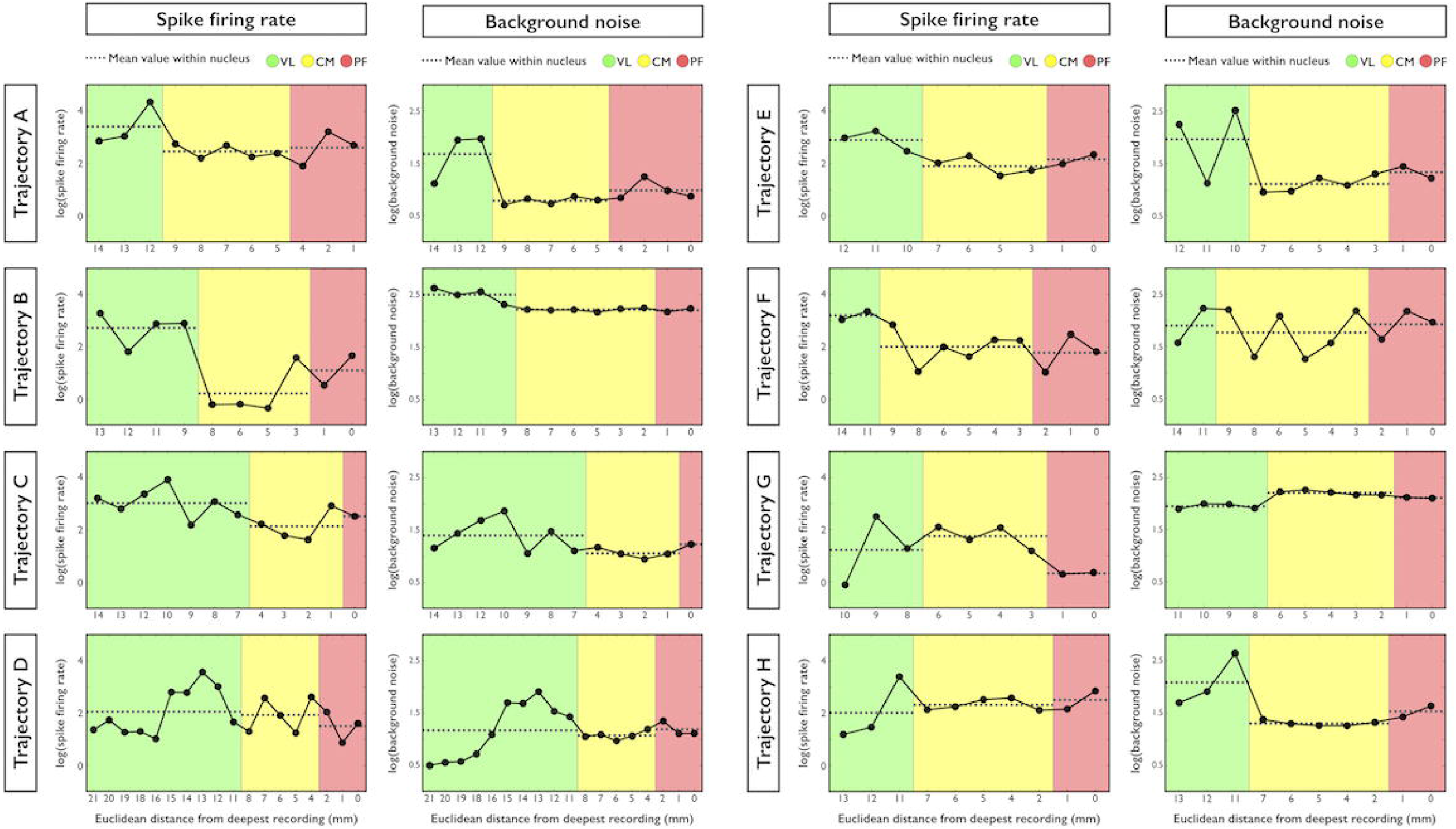
Individual-level analysis of intraoperative microelectrode recordings. Line plots showing spike firing rate and background noise (expressed as log values) from eight individual example microelectrode recording trajectories (one trajectory is shown in each row labelled A to H). The *x* axis of each plot shows the Euclidean distance (mm) between a given recording site and the ‘final’ site (i.e., values on the left of each plot are from more superior thalamic sites, whereas values on the right are from more inferior sites). Recordings are coloured by their thalamic locations defined by the Krauth/Morel atlas[17] (green, yellow, and red areas=recordings within the ventrolateral [VL], centromedian [CM], and parafasicular [PF] nuclei, respectively). The dotted horizontal line within each coloured area indicates the mean value across recordings within the nucleus. ***Interpretation:*** Consistent with the group-level results (Figure 4), most trajectories showed lower mean spike firing rate in CM and PF relative to VL and lower mean background noise in CM relative to VL (e.g., trajectories A-F); however, a minority (20-25%) deviated from the group-level trend in spike firing rate or background noise (e.g., trajectories G and H).

### fMRI connectivity from CM-DBS sites

Figure 7A displays group-level fMRI connectivity calculated from CM-DBS electrode positions. For both left and right sides, positive subcortical connectivity was found with the cerebellum, thalamus, brainstem (midbrain and pons), striatum (caudate and putamen), and sub-thalamic nuclei. Cortically, positive connectivity was prominent in auditory cortex, pre- and postcentral gyri, premotor cortex, cingulate cortex, para-hippocampal/fusiform cortex, and insular cortex. No areas of negative connectivity were found. Spatial similarity with ‘BrainMap’ ICNs (Figure 7B) showed that connectivity was greatest with brain circuits associated with emotional/interoceptive, motor/visuospatial, cerebellar, and auditory/motor speech processing[20 21]. In contrast, connectivity was less apparent with default-mode, fronto-parietal, and visual circuits.

**Figure 7:**
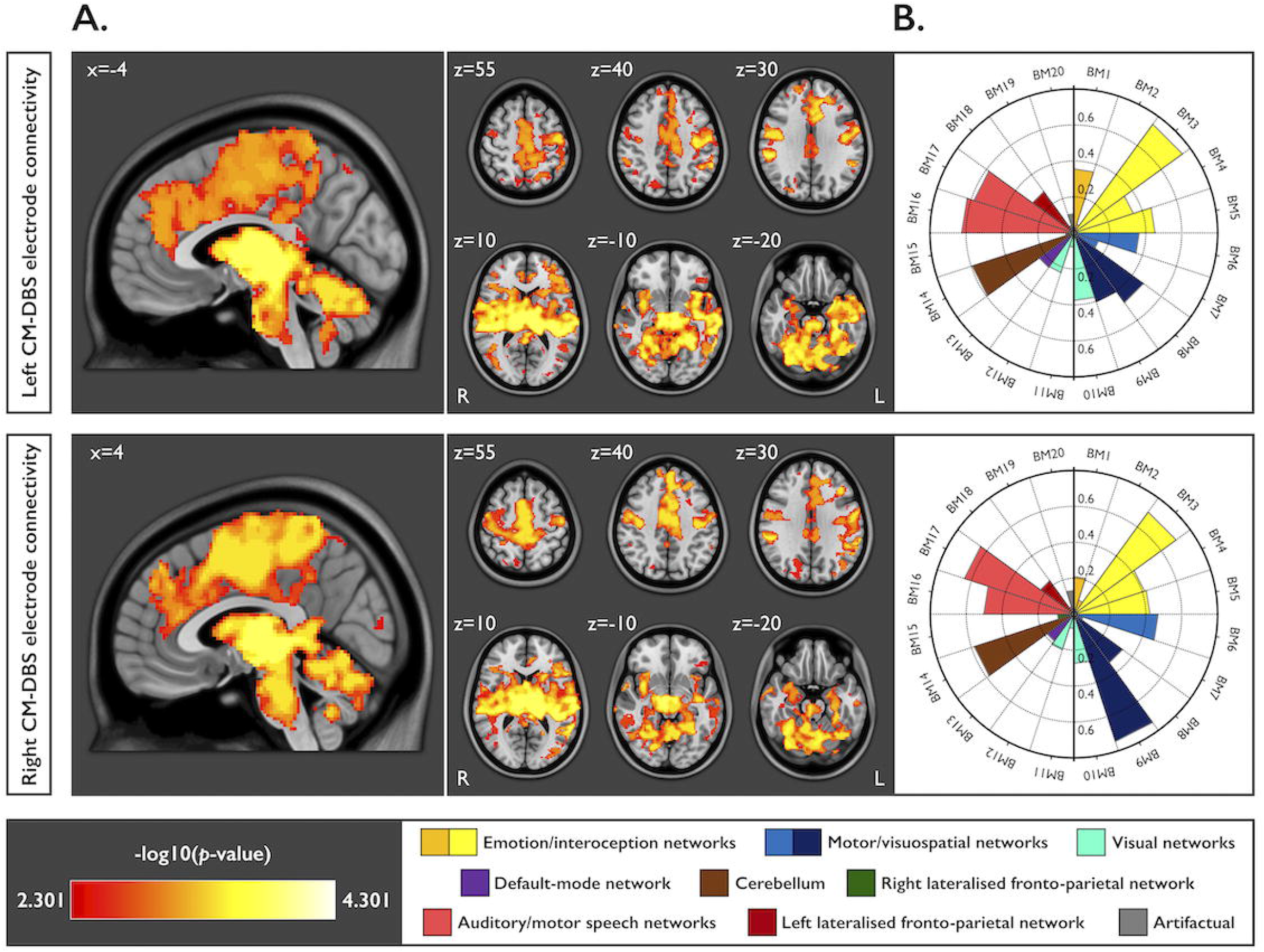
fMRI resting-state functional connectivity from positions of implanted DBS electrode contacts within the centromedian nucleus. **(A)** Sagittal and axial brain maps showing areas of significant positive mean functional connectivity strength (*p*<0.005, corrected for family-wise error after threshold-free cluster enhancement) calculated separately for the left (upper row) and right (lower row) DBS electrode contacts within the centromedian nucleus (CM). Maps are coloured using a -log10 transformation of voxel-wise *p* values. The *x* and *z* coordinates indicate axial and sagittal voxel positions (mm) in Montreal Neurological Institute (MNI)152 2009b space, respectively. **(B)** Polar plots quantifying spatial similarity between the connectivity maps in (A) and each of 20 intrinsic connectivity networks (ICNs) derived from the ‘BrainMap’ (BM) meta-analytic database of >1800 prior task-related neuroimaging studies[20 21]. Spatial similarity is expressed as a value ranging from 0-1 (larger wedges in the plot indicate high similarity with the BM ICN; smaller wedges indicate low similarity). Each wedge is coloured according to the hypothesised functional role(s) of the BM ICN, as indicated by the figure legend in the lower right. ***Interpretation:*** Connectivity of CM-DBS electrode positions is maximal with BM ICNs that normally support emotional/interoceptive, motor/visuospatial, cerebellar, and auditory/motor speech processing; in contrast, connectivity is less apparent with fronto-parietal, default-mode, and visual BM ICNs.

## DISCUSSION

We aimed to provide a comprehensive account of neurosurgical targeting for CM-DBS in the largest clinical trial cohort of LGS patients reported to date. We showed that the CM can be visualised pre-surgically using a novel 3 tesla MP<u>2</u>AGE MRI approach, and that MERs may in some cases provide additional localising features. Using resting-state fMRI connectivity calculated from sites of DBS electrode implantation, we also identified plausible pathways that may mediate therapeutic effects of CM stimulation, including connections with intrinsic brain networks that normally support a range of arousal, cognitive, and sensorimotor processes. In addition to immediate clinical applications in LGS, our findings are relevant to the heterogenous group of neurological and psychiatric diseases for which the CM is considered a potential therapeutic target[1].

The CM’s position on edge-weighted MP<u>2</u>AGE corresponded with multiple sources of atlas and histology information[17 18]. Atlas data suggest the CM lies ∼10mm lateral to the midline and is bordered superiorly by the mediodorsal nucleus, medially by the PF, and posteriorly by the pulvinar (Figure 1). In this location, edge-weighted MP<u>2</u>AGE showed a hyperintense region, presumably the CM, that bordered comparatively hypointense structures consistent with the mediodorsal, PF, and pulvinar nuclei. Visualisation of CM is likely based on the high sensitivity of MP<u>2</u>AGE to intrathalamic variations in grey and white matter (which appear hypo- and hyperintense, respectively)[16], and the greater mix of tissue types within CM relative to adjacent nuclei. This is supported by Nissl preparations of the human thalamus, in which CM is faintly stained and shows a sparse neuronal cell density of ∼2200/mm^3^, whereas mediodorsal, PF, and pulvinar nuclei are darkly stained with higher neuronal densities of ∼3200-5000/mm^3^[25 26].

Few studies have described MRI approaches for identifying CM. At high field strength (7 tesla), both susceptibility-weighted imaging(SWI)[27] and track-density imaging(TDI; derived from diffusion-weighted MRI)[28] have shown potential for visualising multiple thalamic substructures, including CM. Clinical and safety considerations, including anaesthesia requirements and the presence of implanted devices, limited the feasibility of 7 tesla MRI in the patients we studied. Despite this, we acquired SWI and TDI at 3 tesla in a subset of our patients; however, we found the quality of CM visualisation to be less reliable than edge-weighted MP<u>2</u>RAGE (unpublished data). A study using 3 tesla proton density-weighted MRI also reported delineation of the CM in controls[29]; however, the need for coarse slice thickness (∼3mm) to achieve sufficient signal-to-noise ratio may limit the method’s neurosurgical suitability. Functional approaches for localising CM, such as task-based fMRI paradigms that activate CM specifically[30] or thalamic segmentations based on resting-state fMRI connectivity[31], may assist with refining the visualisation achievable by structural MRI; however, further clinical validation is needed.

Prior DBS trials have targeted CM using indirect, stereotactic atlas-based approaches[2 5-9]. The results of our study, which illustrate that indirect targeting could in some cases lead to a gross misplacement of electrodes outside CM (Figure 5), cast some uncertainty on the accuracy of targeting in prior trials. This may contribute to some of the variability reported in the efficacy of CM-DBS for epilepsy, where seizure suppression ranges from 0-100% across different studies[2 5-9]. Given this potential inaccuracy, outcomes from prior studies may not be directly comparable. Hence it is important for future trials to use more direct targeting methods, and to report final electrode positions in a common reference space (e.g. MNI152 2009b space used here; Table 1).

To our knowledge, the utility of MERs for CM-DBS had not been systematically investigated before our study. However, our observation of reduced spike firing rate and background noise in the CM is consistent with a small number of reports in humans[6] and monkeys[32] which describe the posterior intralaminar nuclei as neurophysiologically ‘quieter’ than adjacent structures during spontaneous recordings. One explanation for this finding may relate to the CM’s known roles in selective attention, arousal, and multimodal sensory processing[1], functions that are presumably inhibited during states of reduced awareness such as intraoperative anaesthesia. This is supported by thalamic recordings in patients with disorders of consciousness, where CM spike firing is lower in vegetative/unresponsive patients than those who are minimally conscious[33].

However, some caution should be applied when considering the overall utility of MERs in LGS, given they are not without risks: MERs lengthen surgical time and may increase the frequency of intracranial haemorrhage[34]. Hence they should only be acquired when doing so leads to an unambiguous improvement in accuracy of electrode positioning. Due to inter-patient variability and subtle differences between nuclei in some cases (Figure 6), MERs did not lead to a refinement of the neuroimaging-based target in our study. Their utility was further limited by the possible confounding effects of anaesthesia[35], the impracticality of performing awake recordings in patients with cognitive impairment, and the lack of established methods for evoking spiking patterns specific to the CM (as commonly used in Parkinson’s disease, for example, where passive movement of patients’ limbs alters MER activity within the sub-thalamic nucleus and can thus assist with target identification). Intraoperative stimulations using the microelectrode tip (testing for internal capsule effects)[24] also did not lead to a refinement of our target coordinates. These limitations reinforce the importance of direct targeting methods for LGS.

The pattern of fMRI connectivity calculated from positions of implanted electrodes provides new insights into network mechanisms via which CM-DBS may exert therapeutic effects. Consistent with tracing studies in animals[1 3] and diffusion MRI tractography studies in humans[2], we found connectivity between the CM and the basal ganglia, brainstem, and sensorimotor, premotor, and cingulate cortex. We also found connectivity with additional areas not commonly described in previous studies, including the cerebellum and insular cortex. Although the analysis approach we used does not allow us to infer the CM’s directional or modulatory effects, our results support the view that the CM is connectomically well-positioned to influence networks implicated in a variety of brain diseases[1]. For example, limbic, basal ganglia, and sensorimotor areas are affected in some neuropathic pain syndromes[36], while basal ganglia, cerebellar, limbic, and sensorimotor regions are thought to generate motor and vocal tics in Tourette syndrome[37].

Interestingly, however, the CM’s pattern of cortical connectivity shows some key differences to the pattern of brain activation we previously observed during epileptic activity in LGS. Using simultaneous EEG-fMRI[4] and single-photon emission computed tomography[38], we found prominent activation of fronto-parietal association cortex during tonic seizures and generalised paroxysmal fast activity. We also found that, relative to controls, patients show abnormal fMRI connectivity in the fronto-parietally distributed default-mode and executive-control networks[31]. In contrast, here we found that the CM’s cortical connectivity was greatest with sensorimotor, premotor, and limbic cortex, whereas fronto-parietal connectivity was less apparent.

This result raises the hypothesis that anti-seizure effects of CM-DBS in LGS are not primarily mediated via direct connections with cortical networks through which the epileptic process is maximally expressed. One possibility is that CM-DBS modulates affected cortical areas indirectly, perhaps via dense projections to the dorsal striatum[1], which in turn connects with prefrontal and parietal cortex; or via sparser projections to other thalamic nuclei (e.g., reticular, ventral, and anterior intralaminar nuclei)[3], which likely provide multiple pathways for modulating the cortex more diffusely. This is supported by a previous study in pigs showing that CM-DBS induces fMRI signal inhibition in regions the CM directly projects to (basal ganglia, sensorimotor, and limbic cortex) as well as regions the CM likely influences indirectly, particularly prefrontal cortex[39].

In contrast to LGS, the epileptic network in genetic generalised epilepsy(GGE) involves several areas directly connected to the CM. EEG-fMRI studies[40] show that seizures in GGE prominently activate the striatum and motor/premotor cortex, the CM’s principal efferent targets[1]. This may partly explain the results of a recent CM-DBS trial in 11 patients with a range of epilepsy syndromes, where the four patients with GGE showed the greatest mean seizure suppression[6]. Such knowledge of disease-specific networks may therefore assist with rational selection of patient groups who are most likely to benefit from CM-DBS.

Accurate targeting of the CM for DBS is achievable using a novel ‘edge-weighted MP<u>2</u>RAGE’ MRI approach at clinically available field strength (3 tesla). Intraoperative MERs may in some cases provide neurophysiological indicators of the CM’s location, including reduced spike firing rate and background noise; however, their utility is limited by inter-patient variability and impracticality of non-anaesthetised recordings in cognitively impaired patients. Therapeutic effects of CM-DBS may be mediated via connectivity with multiple intrinsic brain networks normally associated with diverse arousal, cognitive, and sensorimotor functions.

## Data Availability

Data are available on reasonable request.

## ACKNOWLEDGEMENTS

We thank the patients and their families and carers for participating in this research. We also thank the MRI technologists at Austin Health and The Florey Institute of Neuroscience and Mental Health for coordinating scanning, and Amy Weekley and Emily Sewell from Medtronic Australia for assisting with the intraoperative microelectrode recordings. We also acknowledge the facilities and the scientific and technical assistance of the National Imaging Facility at the Florey node, and the support of the Victorian Government through the Operational Infrastructure Support Grant.

## COMPETING INTERESTS

Aaron E.L. Warren, Linda J. Dalic, Annie Roten, Kristian J. Bulluss, and John S. Archer report no competing interests relevant to this study. Wesley Thevathasan has received honoraria from Medtronic and Boston Scientific.

## FUNDING

This study was supported by the National Health and Medical Research Council of Australia (project grant number 1108881) and a seed grant from the Rare Disease Foundation (www.rarediseasefoundation.org) and BC Children’s Hospital Foundation (www.bcchf.ca). Aaron E.L. Warren was supported by a post-doctoral fellowship from the Lennox-Gastaut syndrome Foundation (www.lgsfoundation.org).

## Abbreviations

AC: anterior commissure
DBS: deep brain stimulation
CL: centrolateral nucleus
CM: centromedian nucleus
EEG: electroencephalography
ESTEL: Electrical Stimulation of the Thalamus for Epilepsy of Lennox-Gastaut Phenotype
fMRI: functional MRI
GGE: genetic generalised epilepsy
ICN: intrinsic connectivity network
LGS: Lennox-Gastaut syndrome
MER: microelectrode recording
MNI: Montreal Neurological Institute
MPRAGE: magnetisation-prepared rapid acquisition gradient echo
MP<u>2</u>RAGE: magnetisation-prepared <u>2</u> rapid acquisition gradient echoes
PC: posterior commissure
PF: parafasicular nucleus
ROI: region-of-interest
SWI: susceptibility-weighted imaging
TDI: track-density imaging
VL: ventrolateral nucleus.

## Notes

### Clinical Trial

CT-2016-CTN-05306-1

### Author Declarations

All relevant ethical guidelines have been followed and any necessary IRB and/or ethics committee approvals have been obtained.

Any clinical trials involved have been registered with an ICMJE-approved registry such as ClinicalTrials.gov and the trial ID is included in the manuscript.

## REFERENCES

1. Ilyas A, Pizarro D, Romeo AK, et al. The centromedian nucleus: Anatomy, physiology, and clinical implications. J Clin Neurosci 2019;63:1–7. doi: 10.1016/j.jocn.2019.01.050

2. Kim SH, Lim SC, Yang DW, et al. Thalamo–cortical network underlying deep brain stimulation of centromedian thalamic nuclei in intractable epilepsy: a multimodal imaging analysis. Neuropsychiatr Dis Treat 2017;13:2607–19. doi: 10.2147/NDT.S148617

3. Sadikot AF, Rymar VV. The primate centromedian–parafascicular complex: anatomical organization with a note on neuromodulation. Brain Res Bull 2009;78(2-2):122–30. doi: 10.1016/j.brainresbull.2008.09.016

4. Warren AEL, Harvey AS, Vogrin SJ, et al. The epileptic network of Lennox-Gastaut syndrome: Cortically driven and reproducible across age. Neurology 2019;93(3):e215–e26. doi: 10.1212/WNL.0000000000007775

5. Fisher RS, Uematsu S, Krauss GL, et al. Placebo-controlled pilot study of centromedian thalamic stimulation in treatment of intractable seizures. Epilepsia 1992;33(5):841–51. doi: 10.1111/j.1528-1157.1992.tb02192.x

6. Valentín A, García Navarrete E, Chelvarajah R, et al. Deep brain stimulation of the centromedian thalamic nucleus for the treatment of generalized and frontal epilepsies. Epilepsia 2013;54(10):1823–33. doi: 10.1111/epi.12352

7. Velasco AL, Velasco F, Jiménez F, et al. Neuromodulation of the centromedian thalamic nuclei in the treatment of generalized seizures and the improvement of the quality of life in patients with Lennox–Gastaut syndrome. Epilepsia 2006;47(7):1203–12. doi: 10.1111/j.1528-1167.2006.00593.x

8. Cukiert A, Burattini JA, Cukiert CM, et al. Centro-median stimulation yields additional seizure frequency and attention improvement in patients previously submitted to callosotomy. Seizure 2009;18(8):588–92. doi: 10.1016/j.seizure.2009.06.002

9. Son B-c, Shon YM, Choi J-g, et al. Clinical outcome of patients with deep brain stimulation of the centromedian thalamic nucleus for refractory epilepsy and location of the active contacts. Stereotact Funct Neurosurg 2016;94(3):187–97. doi: 10.1159/000446611

10. Cukiert A, Lehtimäki K. Deep brain stimulation targeting in refractory epilepsy. Epilepsia 2017;58(S1):80–84. doi: 10.1111/epi.13686

11. Schaltenbrand G, Wahren W. Atlas for stereotaxy of the human brain. 2nd ed. Stuttgart: Thieme, 1977.

12. Chan DT, Zhu XL, Yeung JH, et al. Complications of deep brain stimulation: a collective review. Asian J Surg 2009;32(4):258–63. doi: 10.1016/S1015-9584(09)60404-8

13. Goldsmith IL, Zupanc ML, Buchhalter JR. Long-term seizure outcome in 74 patients with Lennox-Gastaut syndrome: Effects of incorporating MRI head imaging in defining the cryptogenic subgroup. Epilepsia 2000;41(4):395–9. doi: 10.1111/j.1528-1157.2000.tb00179.x

14. Wan KR, Maszczyk T, See AAQ, et al. A review on microelectrode recording selection of features for machine learning in deep brain stimulation surgery for Parkinson’s disease. Clin Neurophysiol 2019;130(1):145–54. doi: 10.1016/j.clinph.2018.09.018

15. Möttönen T, Katisko J, Haapasalo J, et al. Defining the anterior nucleus of the thalamus (ANT) as a deep brain stimulation target in refractory epilepsy: delineation using 3 T MRI and intraoperative microelectrode recording. Neuroimage Clin 2015;7:823–9. doi: 10.1016/j.nicl.2015.03.001

16. Marques JP, Gruetter R. New developments and applications of the MP2RAGE sequence-focusing the contrast and high spatial resolution R1 mapping. PloS One 2013;8(7):e69294. doi: 10.1371/journal.pone.0069294

17. Krauth A, Blanc R, Poveda A, et al. A mean three-dimensional atlas of the human thalamus: generation from multiple histological data. Neuroimage 2010;49(3):2053–62. doi: 10.1016/j.neuroimage.2009.10.042

18. Xiao Y, Lau JC, Anderson T, et al. Bridging micro and macro: accurate registration of the BigBrain dataset with the MNI PD25 and ICBM152 atlases. bioRxiv 2019:561118. doi: 10.1101/561118

19. Winkler AM, Ridgway GR, Webster MA, et al. Permutation inference for the general linear model. Neuroimage 2014;92:381–97. doi: 10.1016/j.neuroimage.2014.01.060

20. Kozak LR, van Graan LA, Chaudhary UJ, et al. ICN_Atlas: Automated description and quantification of functional MRI activation patterns in the framework of intrinsic connectivity networks. Neuroimage 2017;163:319–41. doi: 10.1016/j.neuroimage.2017.09.014

21. Laird AR, Fox PM, Eickhoff SB, et al. Behavioral interpretations of intrinsic connectivity networks. J Cogn Neurosci 2011;23(12):4022–37. doi: 10.1162/jocn_a_00077

22. Horn A, Li N, Dembek TA, et al. Lead-DBS v2: Towards a comprehensive pipeline for deep brain stimulation imaging. Neuroimage 2019;184:293–316. doi: 10.1016/j.neuroimage.2018.08.068

23. Dolan K, Martens HC, Schuurman P, et al. Automatic noise-level detection for extra-cellular micro-electrode recordings. Med Biol Eng Comput 2009;47(7):791–800. doi: 10.1007/s11517-009-0494-4

24. Mehanna R, Machado AG, Connett JE, Alsaloum F, Cooper SE. Intraoperative microstimulation predicts outcome of postoperative macrostimulation in subthalamic nucleus deep brain stimulation for Parkinson’s disease. Neuromodulation 2017;20(5):456–63. doi: 10.1111/ner.12553

25. Henderson J, Carpenter K, Cartwright H, et al. Loss of thalamic intralaminar nuclei in progressive supranuclear palsy and Parkinson’s disease: clinical and therapeutic implications. Brain 2000;123(7):1410–21. doi: 10.1093/brain/123.7.1410

26. Highley JR, Walker MA, Crow TJ, et al. Low medial and lateral right pulvinar volumes in schizophrenia: a postmortem study. Am J Psychiatry 2003;160(6):1177–9. doi: 10.1176/appi.ajp.160.6.1177

27. Najdenovska E, Tuleasca C, Jorge J, et al. Comparison of MRI-based automated segmentation methods and functional neurosurgery targeting with direct visualization of the Ventro-intermediate thalamic nucleus at 7T. Sci Rep 2019;9(1):1119. doi: 10.1038/s41598-018-37825-8

28. Calamante F, Oh SH, Tournier JD, et al. Super-resolution track-density imaging of thalamic substructures: comparison with high-resolution anatomical magnetic resonance imaging at 7.0T. Hum Brain Mapp 2013;34(10):2538–48. doi: 10.1002/hbm.22083

29. Kanowski M, Voges J, Tempelmann C. Delineation of the nucleus centre median by proton density weighted magnetic resonance imaging at 3 T. Oper Neurosurg (Hagerstown) 2010;66(Suppl_1):ons-E121-ons-E23. doi: 10.1227/01.NEU.0000348560.85056.63

30. Metzger CD, Eckert U, Steiner J, et al. High field FMRI reveals thalamocortical integration of segregated cognitive and emotional processing in mediodorsal and intralaminar thalamic nuclei. Front Neuroanat 2010;4:138. doi: 10.3389/fnana.2010.00138

31. Warren AEL, Abbott DF, Jackson GD, et al. Thalamocortical functional connectivity in Lennox–Gastaut syndrome is abnormally enhanced in executive-control and default-mode networks. Epilepsia 2017;58(12):2085–97. doi: 10.1111/epi.13932

32. Nanda B, Galvan A, Smith Y, Wichmann T. Effects of stimulation of the centromedian nucleus of the thalamus on the activity of striatal cells in awake rhesus monkeys. Eur J Neurosci 2009;29(3):588–98. doi: 10.1111/j.1460-9568.2008.06598.x

33. Magrassi L, Zippo AG, Azzalin A, et al. Single unit activities recorded in the thalamus and the overlying parietal cortex of subjects affected by disorders of consciousness. PLoS One 2018;13(11):e0205967. doi: 10.1371/journal.pone.0205967

34. Hariz MI. Safety and risk of microelectrode recording in surgery for movement disorders. Stereotact Funct Neurosurg 2002;78(3-4):146–57. doi: 10.1159/000068960

35. Grant R, Gruenbaum SE, Gerrard J. Anaesthesia for deep brain stimulation: a review. Curr Opin Anaesthesiol 2015;28(5):505–10. doi: 10.1097/ACO.0000000000000230

36. Jaggi AS, Singh N. Role of different brain areas in peripheral nerve injury-induced neuropathic pain. Brain Res 2011;1381:187–201. doi: 10.1016/j.brainres.2011.01.002

37. Bohlhalter S, Goldfine A, Matteson S, et al. Neural correlates of tic generation in Tourette syndrome: an event-related functional MRI study. Brain 2006;129(Pt 8):s2029–37. doi: 10.1093/brain/awl050

38. Intusoma U, Abbott DF, Masterton RA, et al. Tonic seizures of Lennox-Gastaut syndrome: Periictal single-photon emission computed tomography suggests a corticopontine network. Epilepsia 2013;54(12):2151–7. doi: 10.1111/epi.12398

39. Kim JP, Min H-K, Knight EJ, et al. Centromedian-parafascicular deep brain stimulation induces differential functional inhibition of the motor, associative, and limbic circuits in large animals. Biol Psychiatry 2013;74(12):917–26. doi: 10.1016/j.biopsych.2013.06.024

40. Masterton RA, Carney PW, Abbott DF, et al. Absence epilepsy subnetworks revealed by event-related independent components analysis of functional magnetic resonance imaging. Epilepsia 2013;54(5):801–8. doi: 10.1111/epi.12163

